# Emerging diseases: when lower vaccine performance in Randomized Clinical Trials means higher economic value

**DOI:** 10.64898/2026.01.19.26344387

**Authors:** Nicolas Houy, Julien Flaig

## Abstract

Using the example of an unknown emerging disease with simple SIR (susceptible-infectious-recovered) dynamics, we show that an efficacy randomized clinical trial (RCT) for a vaccine can be misleading when it comes to the cost-effectiveness of that vaccine. An RCT is more likely to demonstrate efficacy with a high confidence level if it is carried out during the peak of the outbreak. However, in this scenario, the vaccine also has a higher chance of being approved too late to be cost-effective. A vaccine is more likely to be cost-effective if vaccination is implemented in the early stages of an epidemic, but an RCT is more likely to fail to demonstrate efficacy if it is implemented too early, that is when disease transmission is too low.

## 1 Introduction

Cases of COVID-19 were first reported in December 2019 in Wuhan, Hubei Province, China [1]. The first clinical trial of a COVID-19 vaccine (mRNA-1273 by Moderna) began in March 2020 in the United States [2]. By May 2020, 466 clinical trials related to COVID-19 had been identified globally, 35% of which were phase III clinical trials [3]. These figures show that progress in medical science allows, in the case of an emerging disease, the development of vaccines or treatments when the epidemiological characteristics of the disease are barely known or the first phase of the spread of the disease has barely begun [4–7]. A similar situation was witnessed during the 2013-2016 Ebola crisis [8, 9].

The rigorous methodology of clinical trials ensures that only vaccines meeting strict safety and efficacy criteria receive regulatory approval. The final step prior to approval decision is a phase III randomized controlled trial (RCT) whereby the candidate is administered to a large group of volunteers, the treatment group, while another group, the control group, receives a placebo. The candidate’s efficacy is assessed by comparing disease incidence in the treatment and control groups [10, 11]. Vaccine safety is monitored by comparing the incidence of adverse events of interest in the treatment group and in the general population [12, 13]. While ever more advanced methods are being developed for the design and result analysis of RCTs [14], simple efficacy and safety estimands are still used overwhelmingly (see for instance published FDA approval data [15]).

Making a public decision about market access, healthcare coverage, pricing, or reimbursement requires assessing the value of products and intervention, *i.e*. evaluating them based on their costs relative to the health benefits they provide to a target population. This evaluation is performed after and almost always independently of phase III clinical trials [16–18], although merging clinical trials and economic assessment has long been suggested [19–24]. Economic and societal data are typically not considered in the clinical phase [25–27]. Yet in some cases, the information gathered in the clinical phase informs economic actors, public or private. This can be the case when there are “advance market commitments” or other pull mechanisms at work [28, 29], or for company valuation. In all the cited cases, an implicit assumption is made: the economic evaluation will not be in systematic contradiction with clinical evidence - the higher the clinical efficacy, the higher (or the more likely) the social benefit, *and* demonstrated clinical efficacy is a necessary condition for cost-effectiveness.

In the present article, we show with a simple example that this intuition is flawed. Relying on the case of a vaccine against an emerging disease under parameter uncertainty (e.g. a Disease X [30, 31]), we show that phase III clinical trial results can be misleading about the actual benefit of vaccinating the population. The mechanisms underlying this counter-intuitive result can be qualitatively described as follows. Consider the case of an infectious disease emerging in a susceptible population with life-long lasting immunity after infection and basic reproduction number greater than 1. The transmission dynamics of such a disease is well-known. The pathogen spreads as susceptible and infectious individuals come in contact. The number of susceptible individuals decreases as they become infected. The number of infectious individuals reaches a peak and decreases eventually as they recover or die. Over time, transmission slows down and reaches a steady state, possibly without transmission at all, depending on the sustaining vital dynamics.

Now, assume that a vaccine candidate protecting from infection is made available. Assume no side effect for simplicity, and that a phase III clinical trial is implemented to test the candidate’s efficacy. Not considering the dynamics of the disease, similar numbers of infections in the treated and control arms suggest poor vaccine protection, which may support candidate rejection. However, considering the dynamics of the disease, this can also suggest low disease transmission [32]. At the onset of an epidemic, in particular, transmission can be low, but this is precisely when a vaccination campaign can bring the most benefit. On the contrary, demonstrating efficacy with a high confidence level can suggest strong vaccine protection; but it may also be a sign that the RCT was implemented during the peak of the epidemic, and perhaps that is it too late for a vaccination campaign to be cost-effective. Said differently, short-term observations may not reflect plainly the long-term dynamics of an epidemic in general, and especially under uncertainty and in the case of an emerging disease [33–35]. Extrapolating from these short-term observations can be counterproductive.

A wide range of critiques have emerged against the use of p-values in clinical trials. These critiques center around the misinterpretation, misuse, and over-reliance on p-values in making decisions [36–41]. Our critical commentary relies on a different idea. We do not claim that RCT data should be enriched marginally, e.g. with economic or social impact data [27, 42]. Nor do we claim that RCT data should be analyzed differently, by improving existing statistical methods. Instead, we claim that the information brought by this data may be so dependent on the dynamics of the disease as to fail to support decision-making in a systematic way. In our example, if RCT data were to be used to support optimal decision-making, it would have to be interpreted upside-down *i.e*. using efficacy as a signal that it is *not* optimal in expectation to implement a vaccination campaign, and lack of demonstrated efficacy as a signal that it is.

In Section 2, we introduce the model and method we use for our illustrative example. In Section 3, we display the results. Section 4 concludes.

## 2 Material and method

We consider the emergence of an unknown infectious disease in a closed homogeneous population of *N* individuals. Without complete knowledge of the disease, we assume that its spread can be described by a simple and most usual deterministic SIR model with basic reproduction number *R*_0_ and rate of recovery *γ*. Emergence occurs with a single infectious individual at date 0. Later, at date *T*_*v*_, a vaccine candidate is available for a phase III RCT. Each individual to which the vaccine is administered is immediately protected from infection with probability *α*. Vaccine protection does not wane over time. If a vaccination campaign is implemented, all individuals are vaccinated.

Finally, as we will implement a cost-benefit analysis of the vaccine candidate, we consider the following costs:

- For any individual, the daily cost *c*_*i*_ of being infectious. With no loss of generality, we normalize the monetary unit with *c*_*i*_ = 1.
- The vaccination cost per dose *c*_*v*_. This cost covers the vaccine production costs as well as administration costs and possibly adverse events costs.

Notice that we are taking a societal perspective and hence, we do not specify which of the individuals, some institutions, or companies bear the costs. All costs are time discounted with rate *ρ*.

The values of epidemiological parameters *R*_0_, *γ*, and *α* described above are unknown *a priori* to the decision maker. Yet, she has prior beliefs about these parameters taking the form of prior probability distributions. Only *T*_*v*_, the time at which decisions will be implemented, and costs parameters *c*_*i*_ and *c*_*v*_ that we will vary are known without uncertainty. The specific prior distributions for the parameters used throughout this article are given in Table 1.

**Table 1:**
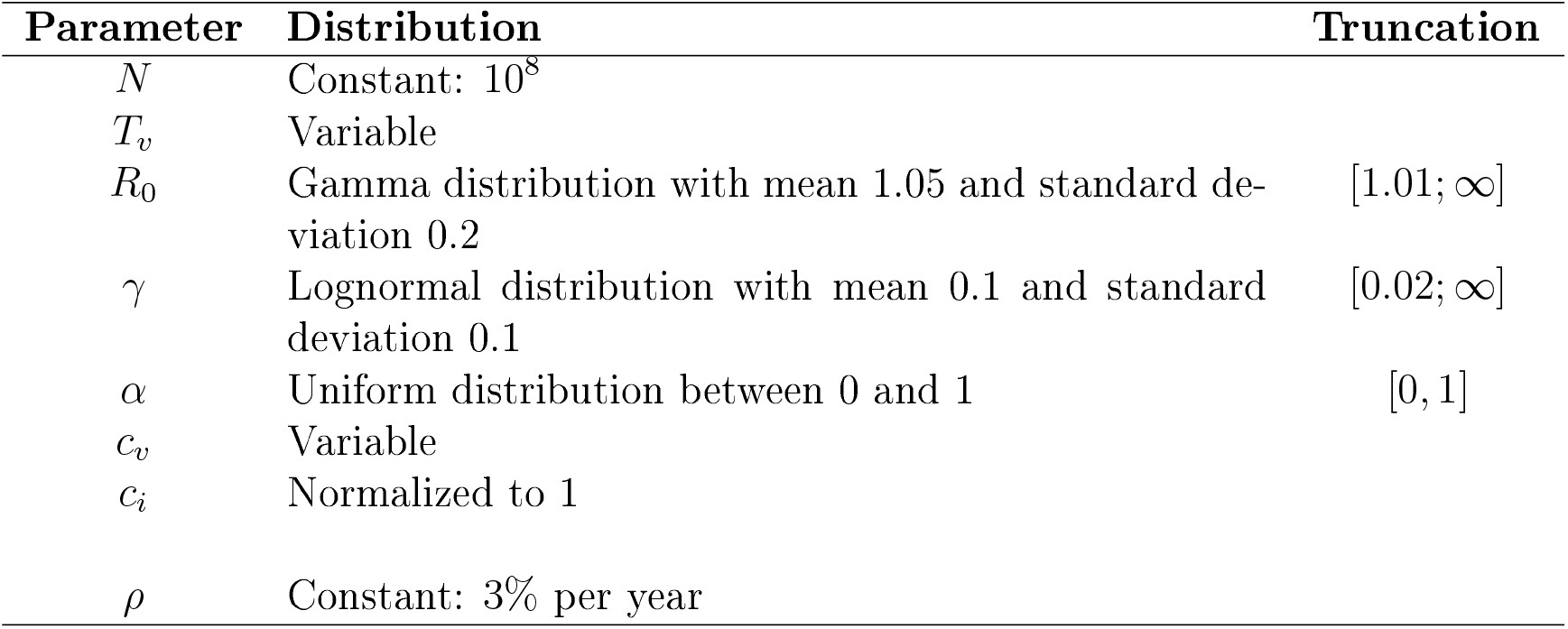
Model parameters prior distributions and values.

In our scenario, an RCT consists in randomly picking a given even number of individuals, *n*, and randomly assigning them to two groups with identical sizes. The individuals of the first group – the control group – remain untreated whereas the individuals in the second group – the treatment group – undergo vaccination. After a given period *T*_*c*_, we compare the number of infections in the control group (*e*_0_) with the number of infections in the treatment group (*e*_1_). We assume Poisson processes for infections in both groups and test the statistical hypothesis that the Poisson rate is smaller for the treated group than for the control group by computing the following z-score: 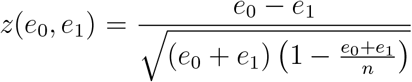, see Casella and Berger [43], Mathews [44], Walpole et al. [45], Devore [46], Triola [47].

## 3 Results

Let us set *T*_*v*_ = 365 days, *T*_*c*_ = 100 days, *n* = 50, 000. We run 100, 000 outbreak and RCT simulations with parameters independently drawn from our prior distributions. The drawn parameter values are displayed in Figure App-1 in Appendix. Table 2 shows the time discounted number of infection-days and number of vaccine doses administered for the following strategies that do not rely on RCT results:

- No vaccination: the RCT is not run and vaccination is never implemented.
- Vaccination at *T*_*v*_: the RCT is not run and vaccination is implemented as soon as vaccine doses are available (at *T*_*v*_).^1^
- Unconditional vaccination after RCT: the RCT is run and vaccination is implemented after the RCT (at *T*_*v*_ + *T*_*c*_), independently of the RCT outcome.
- Unconditional no vaccination after RCT: the RCT is run and vaccination is *not* implemented, independently of the RCT outcome.

**Table 2:** Time discounted number of infection-days (in 10^6^) and administered vaccine doses (in 10^6^) for vaccination strategies not depending on the RCT results.

| Vaccination strategy | Infection-days | Vaccine doses |
| --- | --- | --- |
| No vaccination | 381.15 (95% CI: 378.81 – 383.49) | 0 |
| Vaccination at $T_v$ | 62.02 (95% CI: 61.00 – 63.04) | 97.00 |
| Unconditional vaccination after RCT | 76.94 (95% CI: 75.83 – 78.05) | 96.19 |
| Unconditional no vaccination after RCT | 380.83 (95% CI: 378.49 – 383.18) | 0.02 |

Now, let us investigate how vaccination strategies depending on RCT observations can improve the results. First, let us focus on the usually implemented strategy that consists in vaccinating after RCT if and only if the vaccine efficacy is demonstrated with 95% confidence level (*i.e*. the p-value is below 0.05). Let us denote CT95 this vaccination strategy. With this strategy, the efficacy test is passed with frequency 24.49 % (95% CI: 24.23 – 24.76). The average time discounted number of infection-days is 324.8 (95% CI: 322.5 – 327.2) millions and the average time discounted number of administered vaccine doses is 23.6 (95% CI: 23.3 – 23.8) millions.

Figure 1 shows the monetary gains relative to no vaccination as functions of the vaccination cost, obtained from implementing strategy CT95 as well as the vaccination strategies given in Table 2. Obviously, when vaccination is inexpensive, the best strategy is to vaccinate all individuals as soon as the vaccine is available (vaccination at *T*_*v*_, dotted black line). The line corresponding to unconditional no vaccination after RCT (dashed grey line) is decreasing and close to the x-axis, which it cuts at *c*_*v*_ = 13.03 (not shown). As the cost of vaccination increases, it becomes more profitable to vaccinate less individuals, first only the individuals in the treatment arm of the RCT (unconditional no vaccination after RCT for 3.29 *< c*_*v*_ *<* 13.03), and then not even to implement the RCT at all (*y* = 0 line, not plotted). Interestingly, we can see that there is no vaccination cost for which it is optimal to implement the customary strategy CT95 (plain black line). This does not mean that any clinical trial has no value. However, relying on clinical data to implement vaccination if efficacy is demonstrated with 95% confidence level is never the best strategy in our case, whatever the vaccination cost.

**Figure 1:**
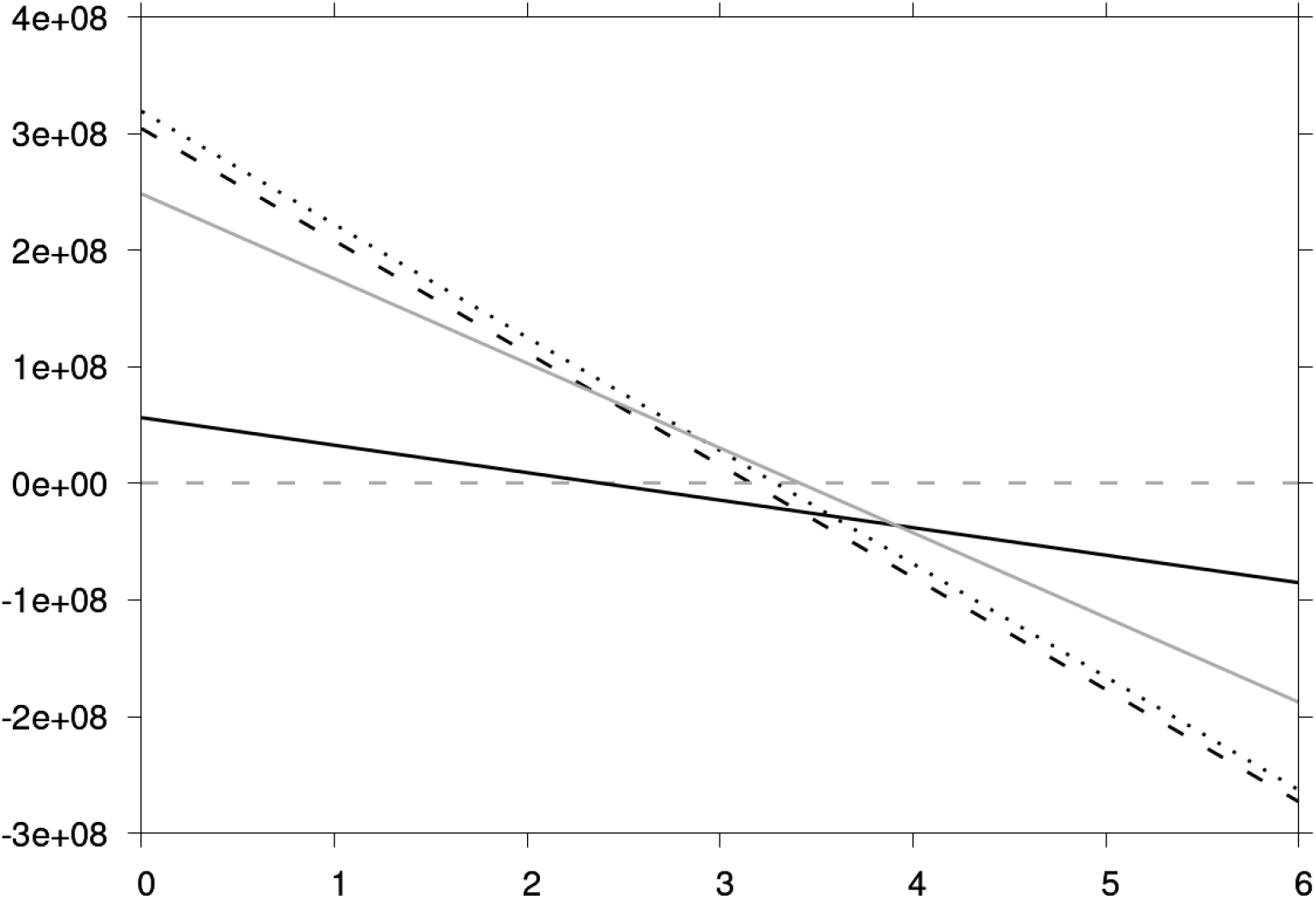
Monetary gains (y-axis) from implementing vaccination at *T*_*v*_ (dotted line), unconditionally implementing vaccination after RCT (dashed black line), unconditionally not implementing vaccination after RCT (dashed grey line, close to *y* = 0), implementing strategy CT95 (plain black line) or implementing strategy 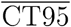 (plain grey line) relative to no vaccination as functions of the vaccination cost (x-axis).

Let us now consider the strategy consisting in vaccinating after the RCT if and only if vaccine efficacy is *not* demonstrated with 95% confidence level. We denote this strategy 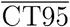. In this case, vaccination is implemented with frequency 75.51 % (95% CI: 75.24 - 75.77) - the set of simulations with vaccine approval under 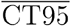 is the complement of those with approval under CT95. The average time discounted number of infection-days is 132.9 (95% CI: 131.4 – 134.5) millions and the average time discounted number of administered vaccine doses is 72.6 (95% CI: 72.4 - 72.9) millions. Notice that this strategy is better than all the strategies considered so far for 2.91 *< c*_*v*_ *<* 3.41 (plain grey line in Figure 1). Also, for *c*_*v*_ *<* 3.91, 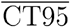 is a better strategy than CT95. Plainly speaking, for this range of vaccination cost, a higher confidence about vaccine efficacy implies a reduced expected benefit of vaccination. Notice also that for 2.38 *< c*_*v*_ *<* 3.41, 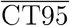 performs better than strategies that implement an RCT but do not use clinical data - unconditional vaccination and unconditional no vaccination after RCT. This means that 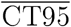 does profitably use the information contained in the clinical data, which we would rather expect from CT95 that does not.

Let us generalize this result to the *whole* set of strategies based on the RCT’s statistical significance. In Figure 2, we display the performance of strategies CT*x* (resp. 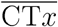) consisting in vaccinating the population if efficacy is demonstrated (resp. not demonstrated) with *x*% confidence level for *x* varying from 0 to 100 in 0.1 increments. In this figure, given vaccination cost *c*_*v*_, all strategies with total cost *C* are located on the line with slope *−*1*/c*_*v*_ and intercept *C/c*_*v*_ (recall that we normalize infection cost *c*_*i*_ to 1). As an illustration, we show in light grey an isocost line with *c*_*v*_ = 3. This isocost is feasible, as it passes through one of the considered strategies. It is also the best among feasible isocosts for *c*_*v*_ = 3, *i.e*. the one with the smallest *C*, as it is the closest to (0,0). This way, we can easily visualize the optimal strategy for any given vaccination cost. We can see that, when compared to the vaccination strategies considered in Figure 2, there is no vaccination cost for which the best strategy is one of the CT*x*’s (black dots). On the contrary, it can be one of the 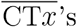 (grey dots). For *c*_*v*_ small enough (isocosts vertical enough), CT*x*’s can be better than 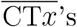, but in this case, unconditional vaccination after RCT (empty triangle) is better than the CT*x*’s. Notice that black dots close to the empty triangle correspond to CT*x*’s with low confidence level and hence almost unconditional vaccination after RCT. Grey dots close to the black triangle correspond to 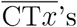 with high confidence level and hence almost unconditional no vaccination after RCT.

**Figure 2:**
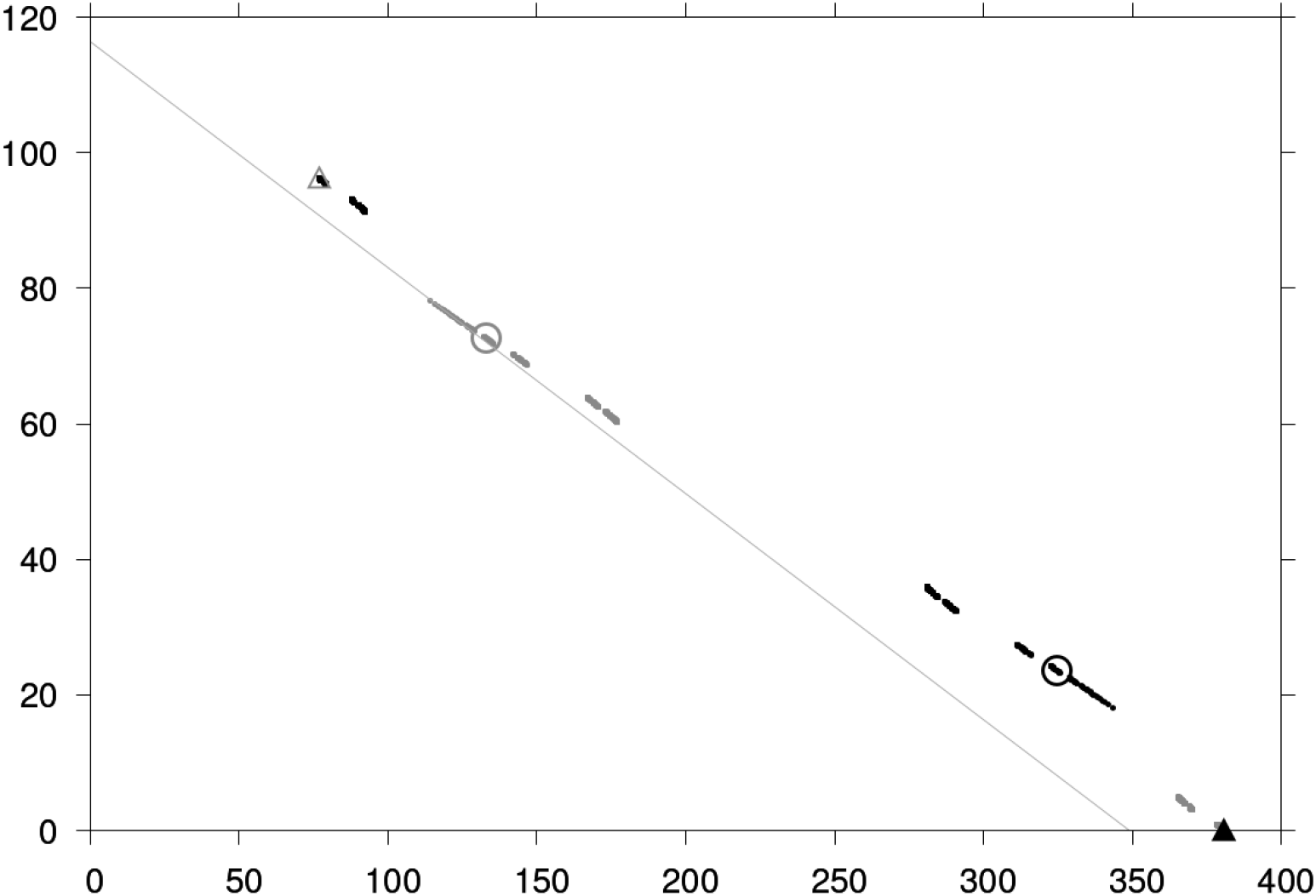
Time discounted number of infection-days (x-axis, 10^6^) and average time discounted number of vaccines administered (y-axis, 10^6^) for the following strategies. Plain triangle: Unconditional no vaccination after RCT; Empty triangle: Unconditional vaccination after RCT; Black points: CT*x*; Grey points: 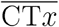; Black circle: CT95; grey circle: 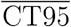.

To sum up, we showed that if we were to use RCT efficacy estimates with high confidence level to decide whether or not to implement a vaccination campaign, then we would have to interpret RCT results in reverse compared to the usual. It seems that the lower the evidence of efficacy, the higher the benefit of a vaccination campaign. Figure 3 illustrates this for *c*_*v*_ = 3. It shows the benefit of a vaccination campaign compared to no intervention after the RCT (the difference in benefit between unconditional vaccination and unconditional no vaccination after RCT) as a function of the RCT’s p-value for each of the 100,000 simulations. The linear regression shows a coefficient of 1.28 (95% CI: 1.18 - 1.37) millions per percentage point in p-value of the RCT. The positive slope of the regression line shows that the statistical significance of the RCT conveys information that comes *against* an incentive to vaccinate.

**Figure 3:**
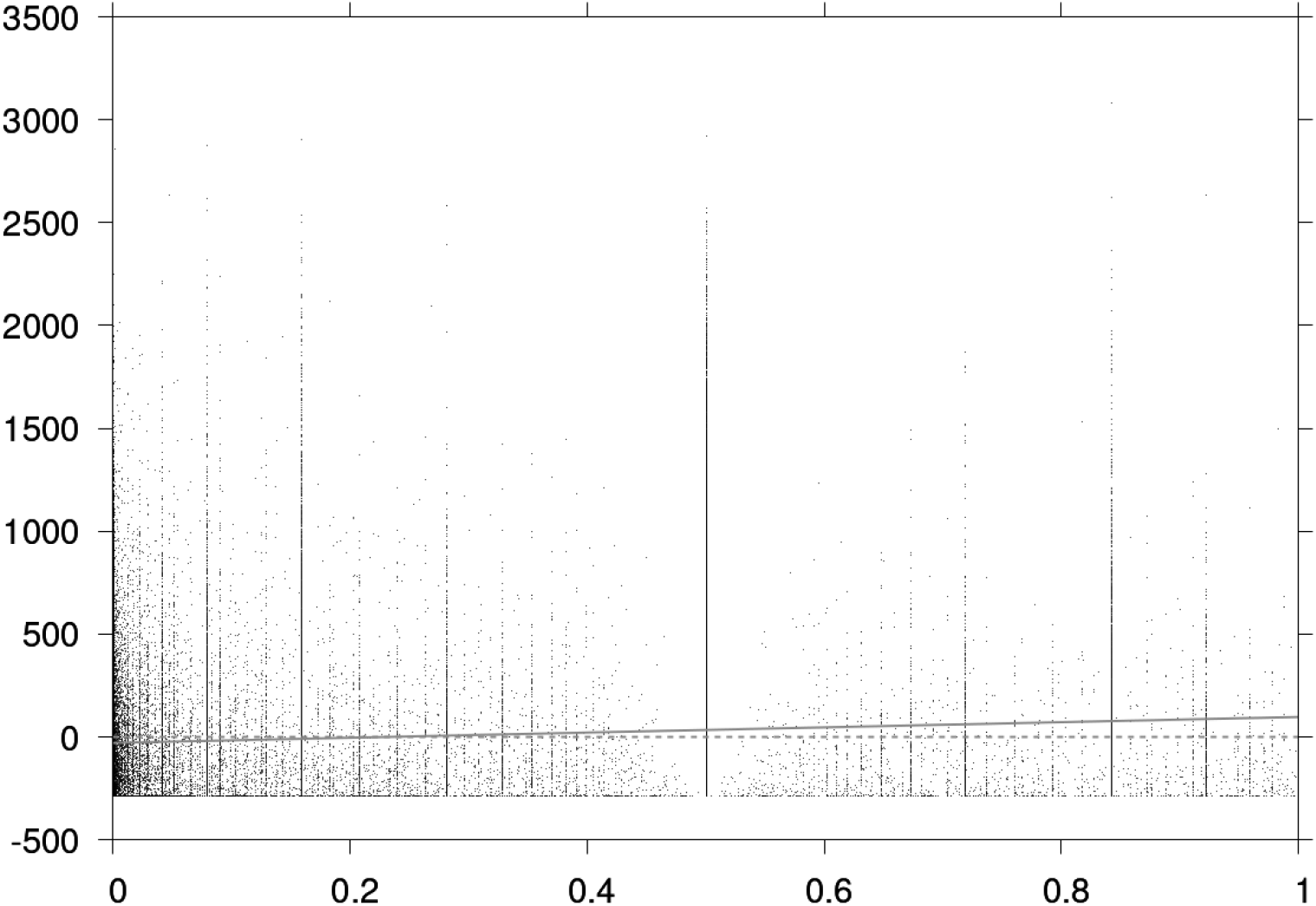
Benefit of a vaccination campaign (in 10^6^) compared to no intervention after the RCT as a function of the RCT’s p-value for each of the 100,000 simulations. Linear regression in light grey.

In particular, let us compare CT95 and 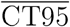. As already said, on average, 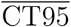 outperforms CT95. The simulation-wise gain to use 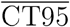 rather than CT95 is 44.72 (95% CI: 42.50 – 46.95) millions.^2^ The difference holds for each of the two decisions - approve or reject the vaccine candidate. Indeed, for the simulations in which the candidate is rejected based on CT95’s efficacy test (hence, by definition, simulations in which it is accepted based on 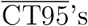 efficacy test), the benefit of a vaccination campaign after RCT (*i.e*. of following 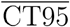 instead of CT95) is 39.80 (95% CI: 37.11 – 42.49) millions. And for simulations in which the candidate passes CT95’s efficacy test, the benefit of a vaccination campaign after RCT is -59.89 (95% CI: -63.64 - -56.15) millions *i.e*. it is better to follow 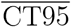 and to not vaccinate. Hence, 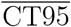 is better than CT95 at *both* approving and rejecting candidates.

Let us now explain this counter-intuitive result. We call unmitigated peak time (UPT) the date at which the number of infectious individuals is the highest in the absence of intervention. Let us split simulations into two subsets: those for which the UPT is below two years and those for which the UPT is above two years. Figure 4 shows the same linear regressions as Figure 3 for each subset. Linear regressions display negative coefficients: - 2.83 (95% CI: -2.95 – -2.71) millions per percentage point in RCT’s p-value for simulations with UPT below two years (27,751 simulations), and -1.64 (95% CI: -1.79 – -1.48) millions per percentage point RCT’s p-value for simulations with UPT above two years (72,249 simulations). Recall that the linear regression using the full set of simulations, by contrast, has a positive coefficient.^3^

**Figure 4:**
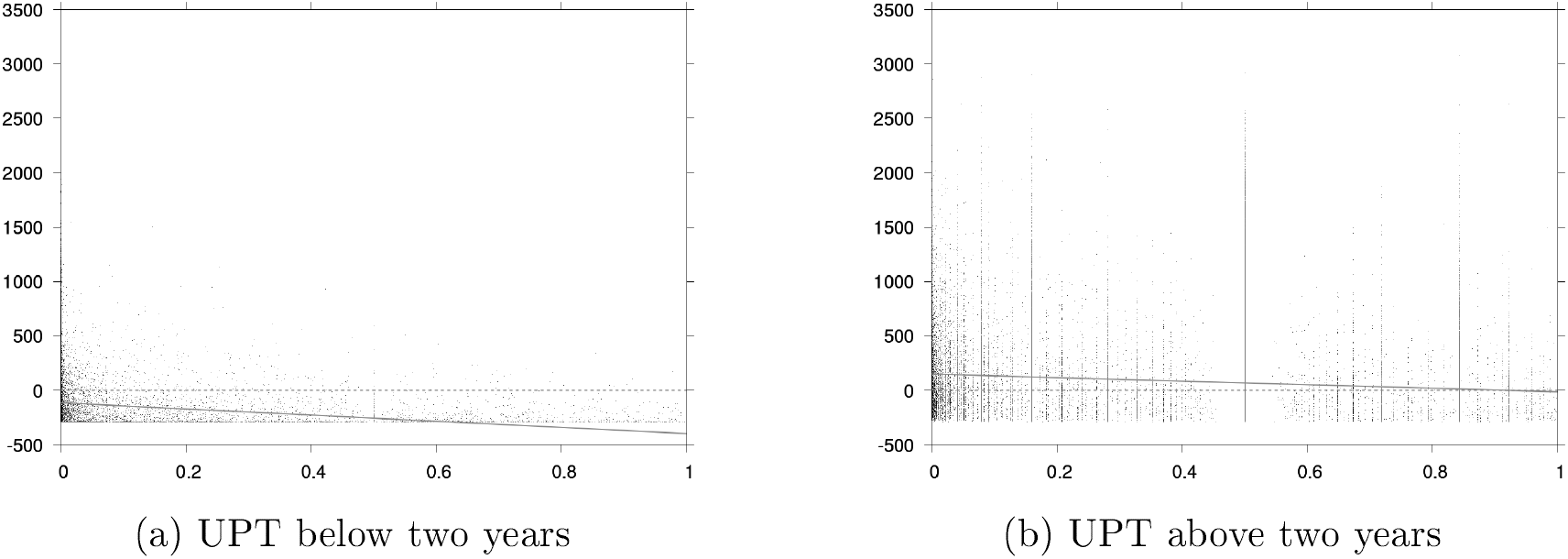
Benefit of a vaccination campaign (in 10^6^) compared to no intervention after the RCT as a function of the RCT’s p-value for each of the 100,000 simulations in two subsets: unmitigated peak time (UPT) below and above two years. Linear regressions in light grey.

Intuitively, the higher the efficacy estimate (the smaller the p-value), the more likely vaccination will be cost-effective in each subset. This intuition relies on the idea that RCT data conveys information about the inherent properties of the vaccine. However, this same intuition is flawed for the whole set of simulations. Indeed, RCT data also conveys information about the disease dynamics, which in turns influences cost-effectiveness (*i.e*. disease dynamics is a confounder).

Consider the subset of simulations with UPT below two years (27.75% of the simulations). These simulations display comparatively faster disease dynamics. Under CT95, the candidate is approved in 72.78% of the simulations. However, it is not optimal to vaccinate: the average benefit of vaccination is negative and equal to -110.0 (95% CI: -113.4 – -106.6) millions for the simulations in which the candidate is approved, and to -245.2 (95% CI: - 247.9 – -242.4) millions for the simulations in which the candidate is rejected. Now, consider the subset of simulations with UPT above two years (72.25% of the simulations). These simulations display comparatively slower disease dynamics. Under CT95, the candidate is approved in only 5.95% of the simulations. However, it is optimal to vaccinate: the average benefit of vaccination is positive and equal to 175.6 (95% CI: 163.9 – 187.3) millions for the simulations in which the candidate is approved, and to 71.5 (95% CI: 68.6 – 74.4) millions for the simulations in which the candidate is rejected.

With faster disease dynamics, the RCT is more likely to demonstrate efficacy and the candidate to be approved, but the vaccination campaign is also more likely to be implemented too late to be cost-effective.^4^ With slower disease dynamics, the RCT is more likely to fail to demonstrate efficacy and the candidate to be rejected, but a vaccination campaign is more likely to be cost-effective. From the perspective of decision-making, a successful RCT likely indicates that it is already too late to implement a vaccination campaign, while failing to demonstrate efficacy likely indicates that it is still time. These ideas are illustrated in Figure 5. In Figure 5a (resp. 5b), we show the number of infectious individuals without vaccination after the RCT (black curve) or with vaccination after the RCT (grey curve) as functions of time when the disease dynamics has an early (resp. late) UPT.^5^ The number of infectious days averted by vaccination is the area between the grey and black curves. It can be seen that an early UPT corresponds to a small vaccination benefit. In Figures 5c and 5d, we show the cumulative incidence for the same dynamics and vaccination policies as in Figures 5a and 5b. If the cumulative incidence during the RCT (denoted *A*) is too small, then the confidence in the vaccine efficacy will likely be small. This occurs in the late UPT case.

**Figure 5:**
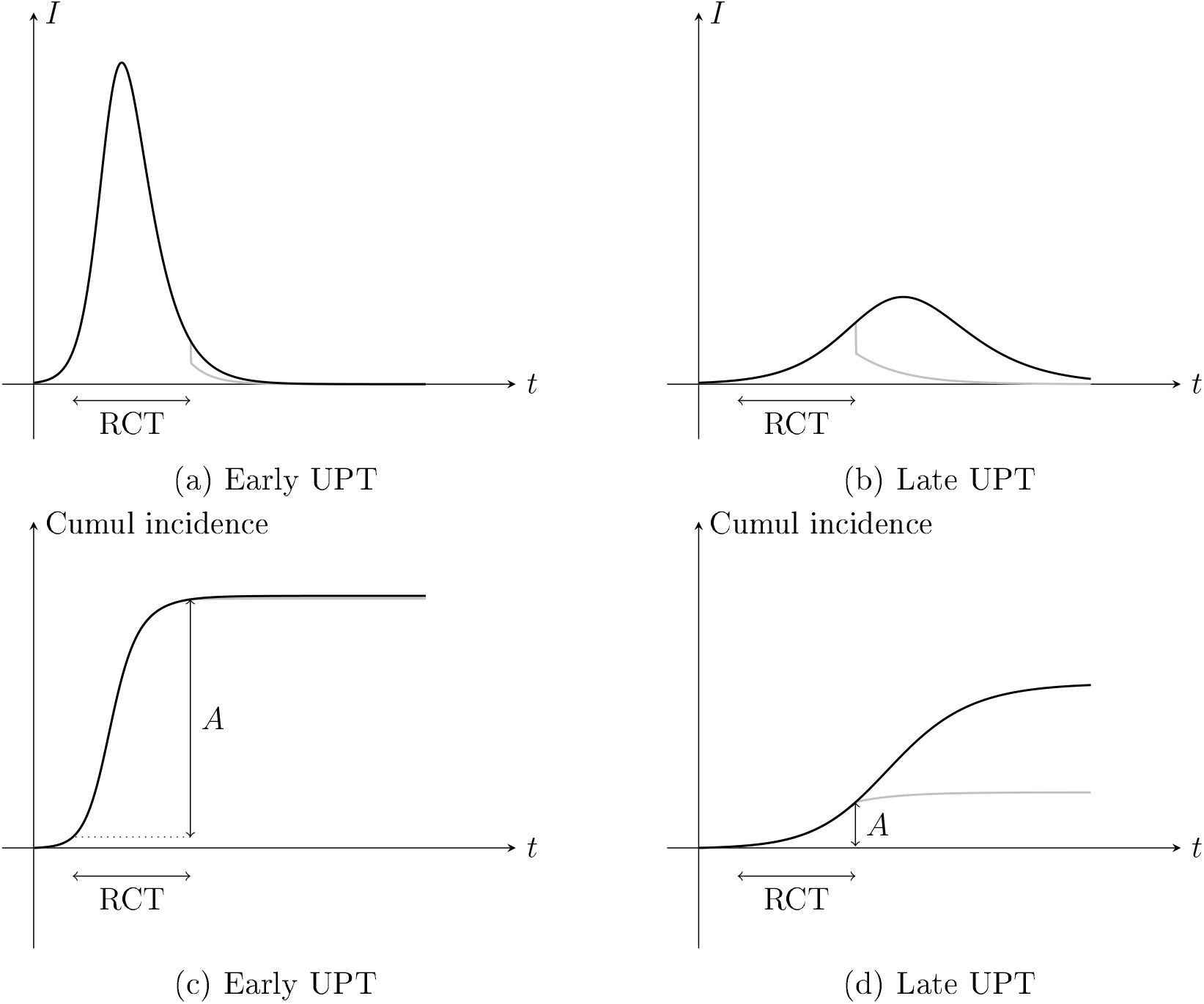
Top: number of infectious individuals without vaccination after the RCT (black curve) or with vaccination after the RCT (grey curve) as functions of time when the disease dynamics has an early (left) or late (right) UPT. Bottom: cumulative incidence without vaccination after the RCT (black curve) or with vaccination after the RCT (grey curve) as functions of time when the disease dynamics has an early (left) or late (right) UPT. *A* denotes the cumulative incidence during the RCT.

## 4 Conclusion

In this article, we showed, using an illustrative example, how RCT efficacy data can be misleading about the health-economic or public health value of a vaccine. In our scenario, the dynamics of the disease and parametric uncertainties are critical. Indeed, RCT data is influenced by the stage of the epidemic: demonstrating efficacy with a high confidence level means that a large enough number of cases was reached in the trial, which indicates a late epidemic stage and, as a consequence, a lower incentive to vaccinate. Uncertain parameters influence both the RCT and the cost-effectiveness of vaccination and make it difficult to extrapolate from short-term observations.

With this article and this stylized example, we add to the numerous critics of phase III clinical trials as they stand. Relying on RCT data to support decision-making usually assumes implicitly that statistical confidence correlates positively with health-economic or public health value, and that statistically demonstrated clinical efficacy is a prerequisite for positive impact. We showed that these assumptions must be questioned in situations in which the disease dynamics and uncertainties are important features. The practical difficulty of implementing RCTs in such situations is well documented. We go further and show that even assuming that an RCT can be implemented technically, its conclusions can be deeply misleading. This can be the case in disease emergence scenarios, but we could generalize to seasonal epidemics or the emergence of new variants.

One could convincingly argue that phase III clinical trials are not about health-economic or public health value. It could be argued that targeting a given level of efficacy is justified by compliance with an ethical imperative. Our article should be read as a proof that there can be important social costs associated with implementing this ethical imperative. These costs are two-fold: in our example, phase III clinical trials systematically tend to prevent vaccines to be used even though their social benefit would be positive, and they favor vaccines for which the social benefit is negative.

## Data Availability

All data produced in the present study are available upon reasonable request to the authors

## A Additional tables and figures

**Figure App-1:**
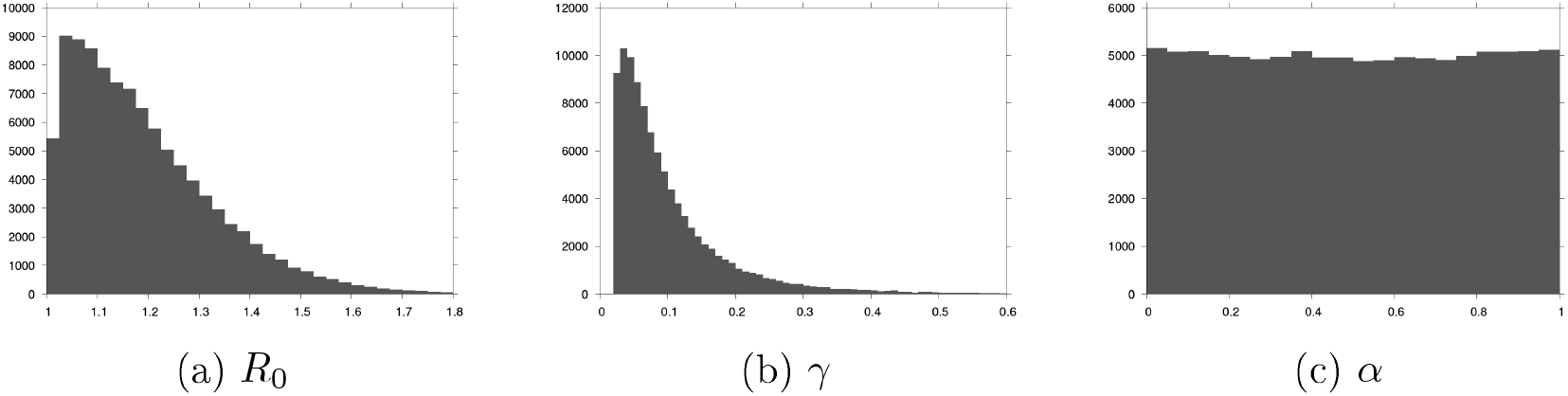
Distribution of the values of the parameters for the 100,000 simulations.

**Table App-1:** Depending on the vaccination strategy, ratio of simulations for which:

| Vaccination strategy | TP | FP | FN | TN |
| --- | --- | --- | --- | --- |
| Unconditional vaccination after RCT | 36.4% | 63.6% | 0.0 | 0.0% |
| Unconditional no vaccination after RCT | 0.0 | 0.0% | 36.4% | 63.6% |
| CT95 | 7.0% | 17.4% | 29.4% | 46.1% |
| $\overline{\text{CT95}}$ | 29.4% | 46.1% | 7.0% | 17.4% |

- TP: vaccination is implemented and it is optimal to implement vaccination after RCT.
- FP: vaccination is implemented and it is optimal not to implement vaccination after RCT.
- FN: vaccination is not implemented and it is optimal to implement vaccination after RCT.
- TN: vaccination is not implemented and it is optimal not to implement vaccination after RCT.

**Table App-2:**
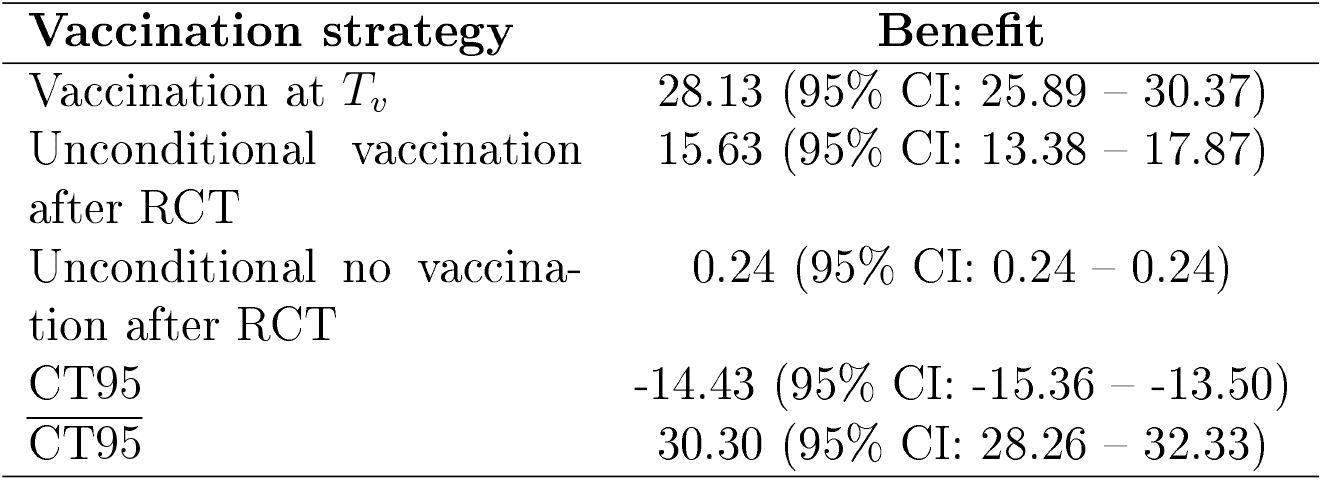
Benefit from different vaccination strategies relative to no intervention for *c*_*v*_ = 3 (in 10^6^).

**Table App-3:**
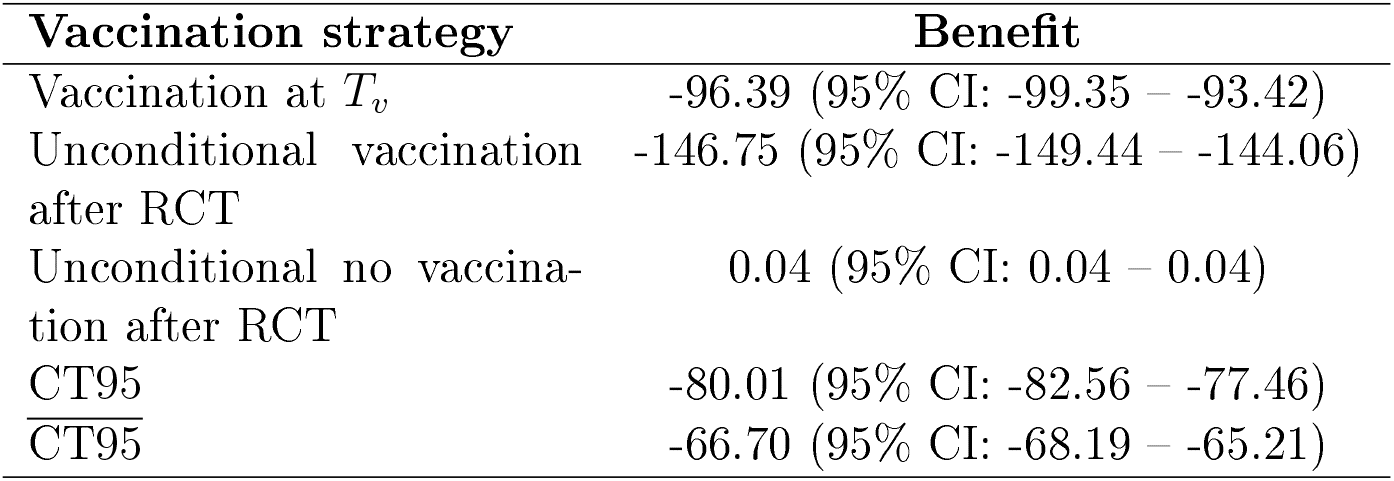
Benefit from different vaccination strategies relative to no intervention for *c*_*v*_ = 3 (in 10^6^) and for simulations with UPT below two years (27,751 data).

**Table App-4:**
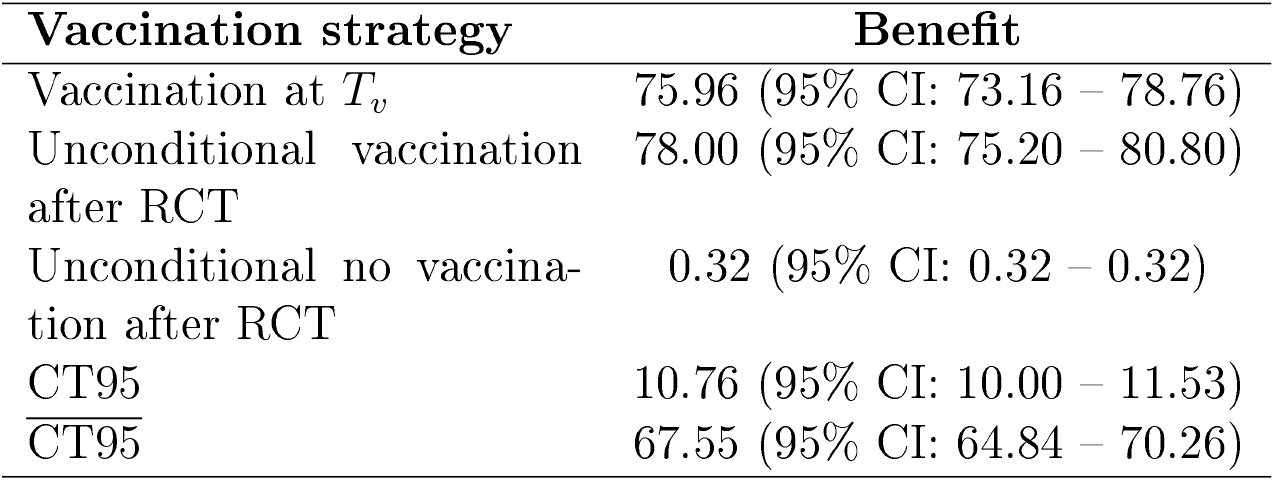
Benefit from different vaccination strategies relative to no intervention for *c*_*v*_ = 3 (in 10^6^) and for simulations with UPT above two years (72,249 data).

**Figure App-2:**
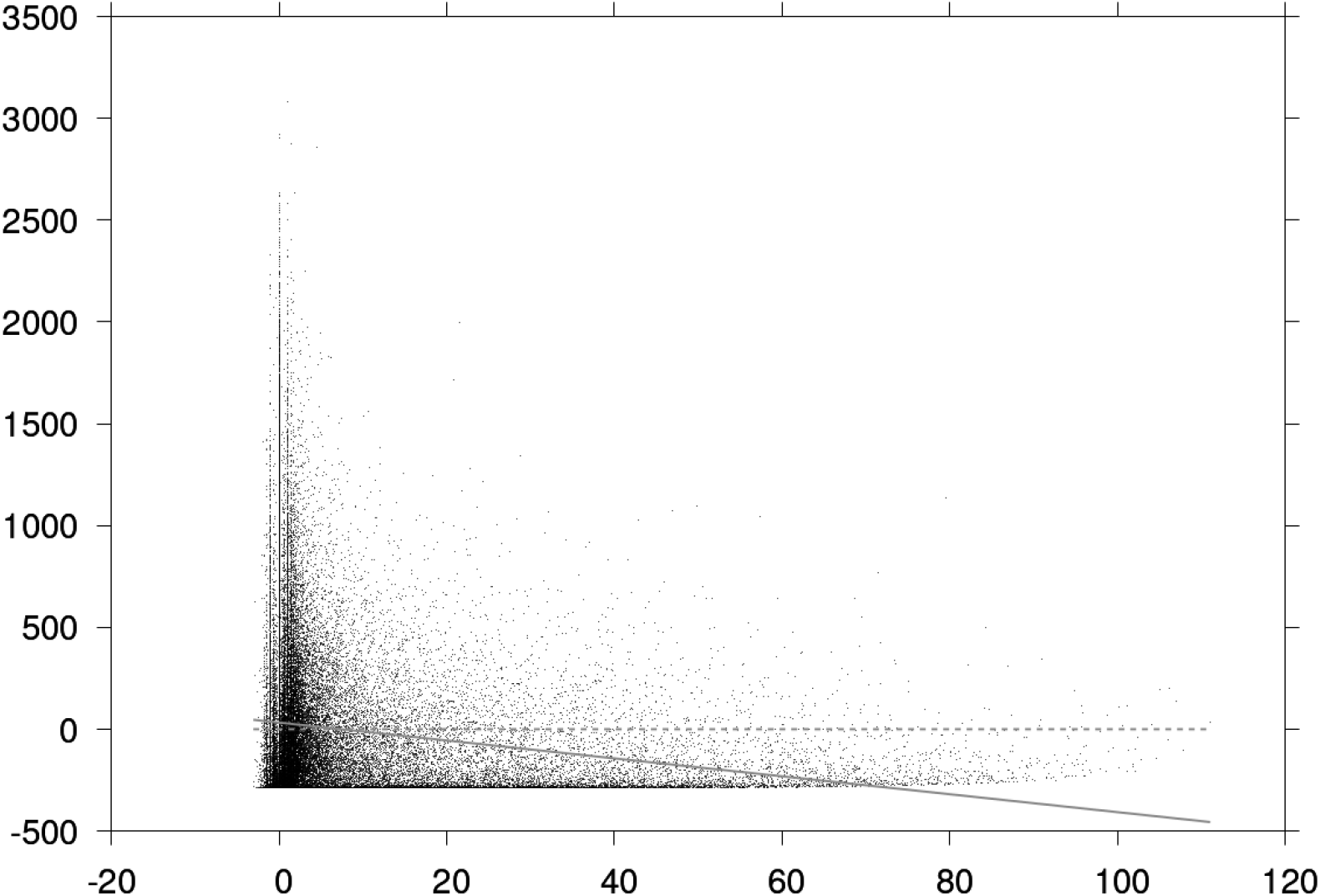
Benefit of a vaccination campaign (in 10^6^) compared to no intervention as a function of the RCT’s p-value for each of the 100,000 simulations. Linear regression in light grey.

## Footnotes

1 We consider that vaccine doses are immediately available and administered without any production or logistic delay.

2 Absolute values are summed up in Table App-2.

3 This is an example of Simpson’s paradox 48 .

4 Notice that for an even faster dynamics, the number of recovered is more important and hence, again, the RCT is less likely to demonstrate efficacy.

5 All dots in Figures 5a and 5b correspond to dots in Figure 3.

